# SARS-CoV-2 infections in people with PCD: neither frequent, nor particularly severe

**DOI:** 10.1101/2020.12.20.20248420

**Authors:** Eva SL Pedersen, Myrofora Goutaki, Amanda L Harris, Lucy Dixon, Michele Manion, Bernhard Rindlisbacher, COVID-PCD patient advisory group, Jane S Lucas, Claudia E Kuehni

## Abstract

People with pre-existing chronic health conditions are reportedly at high risk of getting the coronavirus disease (COVID-19) and of having a severe disease course but little data exist on rare diseases such as Primary Ciliary Dyskinesia (PCD). We studied risk and severity of SARS-CoV-2 infections among people with PCD using data from the COVID-PCD, a participatory study that collects data in real-time directly from people with PCD. Data was collected using online questionnaires. A baseline questionnaire collected information on demographic data, information about the PCD diagnosis and severity. A short weekly questionnaire collected information about current symptoms and incident SARS-CoV-2 infections. 578 people participated in the COVID-PCD by December 7, 2020, with a median number of follow-up weeks of 9 (interquartile range: 4-19 weeks). 256 (45%) of the participants had been tested for SARS-CoV-2 and 12 tested positive prior to study entry or during study follow up (2.1% of the total included population, 95% confidence interval (CI) 1.1-3.6%). 4 people tested positive during the study follow-up, corresponding to an incidence rate of 2.5 per 100 person-years (95% CI: 0.9-6.5). Overall, reported severity was mild with two reporting no symptoms, eight reporting mild symptoms, one reporting severe symptom without hospitalisation, and one reporting hospitalisation for 9 days. The study suggests that with careful personal protection, people with PCD do not seem to have an increased risk of infection with SARS-COV-2, nor an especially severe disease course.

**Take home message:** In this longitudinal study of people with PCD followed weekly via online questionnaires, the incidence rate of COVID-19 and the proportion of participants infected were low, and the observed severity mostly mild.

---

The coronavirus disease 2019 (COVID-19) pandemic caused by SARS-CoV-2 has by December 2020 infected at least 70 million people worldwide and caused over 1.5 million deaths. People with pre-existing chronic health conditions are reportedly at high risk of catching the disease and of having a severe disease course [1-4]. Primary Ciliary Dyskinesia (PCD) is a multisystem, genetic disease which affects about 1 in 10,000 people and leads to chronic upper and lower airway disease, laterality defects, including congenital heart disease, and other health problems [5-8]. In March 2020, PCD patient support groups contacted the paediatric respiratory research group in the University of Bern with the wish to set up a study that generates evidence on the risk and evolution of COVID-19 in people with PCD. This led to the launch of COVID-PCD, a longitudinal online survey of health, shielding behaviours, and quality of life of people with PCD during the pandemic. COVID-PCD is a participatory study that collects data in real-time directly from people with PCD using online questionnaires. This manuscript provides first data on risk and severity of SARS-CoV-2 infections among study participants for the time period between May 30^th^ and December 7^th^ 2020.

A detailed description of the methods has been published [9]. In short, COVID-PCD is an international study advertised through PCD support groups and is open to people of any age with a confirmed or suspected diagnosis of PCD who can complete questionnaires in English, German, or Spanish. French and Italian versions will soon be available. The study has been approved by the cantonal ethics committee of Bern (Study ID: 2020-00830), is registered with clinical trials gov (NCT04602481), and is anonymous. Since May 30^th^ 2020, participants can register and consent via the study website (www.covid19pcd.ispm.ch) and then receive e-mail links to online questionnaires. A baseline questionnaire collects demographic data, information about the PCD diagnosis and severity using the standardised FOLLOW-PCD questionnaires [10], and information on SARS-CoV-2 infections that had occurred prior to study entry. One week after completing the baseline questionnaire, and in weekly intervals thereafter, participants receive short follow-up questionnaires about current symptoms, shielding behaviour, and incident SARS-CoV-2 infections. Questions asking about incident SARS-CoV-2 infections refer to the time passed since completing the last follow-up questionnaire, ensuring that all incident SARS-CoV-2 infections are reported, even if a participant fails to complete a weekly questionnaire. Parents complete questionnaires for children under age 13 years.

We described the number and proportion of study participants who had received a test for SARS-CoV-2 at any time, summing up antigen tests and antibody tests. We then calculated the proportion of people with a confirmed SARS-CoV-2 infection by dividing the number of those with a positive PCR or antibody test at any time (prior to study entry, or during the observation period) by the study population. Participants were asked how seriously ill they got, with answers categorised as no symptoms, mild symptoms (mild fever or cough), moderate symptoms (high fever and cough or dyspnoea, but not hospitalized), severe symptoms resulting in a hospitalisation and very severe disease resulting in intensive care, artificial ventilation, or death. We also calculated the incidence rate of SARS-CoV-2 infections in those who had been disease-free at study entry (the population at risk). We defined an incident case as a positive SARS-CoV-2 test result reported at least 14 days after study entry. This criterion was set to minimize the risk of selection bias from people registering because of typical symptoms or contact with a case. We defined person-time at risk as time between completing the baseline questionnaire and the latest follow-up among those without SARS-CoV-2 at baseline, assuming no re-infections. For each observation week, we calculated the proportion of study participants who reported behaviours related to shielding such as not leaving the house, visiting grocery stores, going to school or work, and using public transport and then averaged these proportions over all observation weeks.

By December 7^th^ 2020, 578 persons with PCD had registered in COVID-PCD, including 219 young people aged less than 19 years and 359 adults aged 20 years or more **(table 1)**. Sixty-one percent were female. The longest time a participant was followed up was 26 weeks (median number of follow-up weeks: 9, interquartile range: 4-19 weeks). 319 participants (55%) had never been tested for SARS-CoV-2, 146 (25%) had been tested once, and 113 (20%) twice or more. Twelve participants reported a positive SARS-CoV-2 test either at study entry or during the observation period, corresponding to 2.1% of the study population (95% CI: 1.1-3.6%). Four cases occurred in those aged under 20, and 8 in people age 20 years or more. Overall, reported severity in the 12 cases was mild, with two reporting no symptoms, eight reporting mild symptoms, one reporting moderate symptoms without hospitalization, and one person (aged 46 years) reporting severe symptoms resulting in a hospitalizing for 9 days. None needed intensive care or artificial ventilation, and none died from COVID-19. Eight of the 12 infections were reported at the time of registration into the study, and 4 incident infections were observed during follow-up. The total follow-up time was 58,805 days (161 person-years). This resulted in an incidence rate of 2.5 per 100 person years (95% CI 0.9-6.5) – meaning that if 100 participants had been observed for a year, two and a half would have caught COVID-19 during this year. Incidence was roughly similar in children with 3.1 per 100 person-years (95% CI 0.8-12.2) and adults with 2.1 per 100 person years (95% CI 0.5-8.1). During the follow-up period, 10% of study participants on average reported not to have left their house during the last 7 days (range: 3 to 17%). 44% had left the house for grocery shopping in the past week (range: 22-46%), 43% had been to school or workplace (range: 16-57%), and 15% had used public transportation (range: 5 to 18%). These proportions varied from week to week and between regions.

**Table 1:** **SARS-CoV-2 infections and shielding behaviour in people with Primary Ciliary Dyskinesia**, based on longitudinal data from the COVID-PCD study

|  | **Total**<br>N=578<br>n (%) | **Children**<br>(0-19 years)<br>N=219<br>n (%) | **Adults**<br>(≥20 years)<br>N=359<br>n (%) |
| --- | --- | --- | --- |
| Male | 221 (39) | 113 (52) | 110 (31) |
| Female | 352 (61) | 106 (48) | 247 (69) |
| **Tested for SARS-CoV-2** |  |  |  |
| never | 319 (55) | 123 (53) | 196 (55) |
| once | 146 (25) | 61 (28) | 85 (24) |
| twice or more | 113 (20) | 35 (16) | 78 (22) |
| **Confirmed SARS-CoV-2 infections** (positive PCR or antibody test at any time during study period, N (%), 95% CI)) | **12 (2.1, 1.1-3.6)** | **4 (1.8, 0.5-4.6)** | **8 (2.2, 1.0-4.3)** |
| No symptoms | 2 (17) | 2 | 0 |
| Mild symptoms, not hospitalised <sup>a</sup> | 8 (67) | 1 | 7 |
| Moderate symptoms, not hospitalised <sup>b</sup> | 1 (8) | 1 | 0 |
| Severe symptoms, hospitalised <sup>c</sup> | 1 (8) | 0 | 1 |
| Very severe symptoms (ICU care, intubation or death) | 0 | 0 | 0 |
| **Incident infections** reported during follow-up period | 4 | 2 | 2 |
| Total follow-up time (in person-years) | 161 | 64 | 97 |
| Incidence rate (infections per 100 person years (95% CI)) | 2.5 (0.9-6.5) | 3.1 (0.8-12.2) | 2.1 (0.5-8.1) |
| **Shielding behaviour during the last 7 days** (mean proportion (range) per week <sup>d</sup> during 25 follow-up weeks)) |  |  |  |
| Did not leave the house | 10% (3-17) | 9% (1-16) | 10% (3-19) |
| Went for grocery shopping | 44% (22-46) | 20% (6-26) | 58% (24-66) |
| Went to workplace/school <sup>e</sup> | 43% (16-57) | 48% (16-75) | 37% (9-46) |
| Used public transportation | 15% (5-18) | 15% (0-20) | 16% (8-20) |

<sup>a</sup>Referred to in the questionnaire as "mild fever or cough" <sup>b</sup>Referred to in the questionnaire as "high fever, cough, headache, etc." <sup>c</sup>The person treated at hospital was not treated in the intensive care ICU Intensive care unit <sup>d</sup>we calculated the proportion of people in each week, and then averaged this proportion over the 25 weeks of the observation period.
<sup>e</sup>Among those who go to school or work.

In summary, this international longitudinal study of nearly 600 people with PCD found that only 2.1% of the study population had had a SARS-CoV-2 infection confirmed by a specific test. This was lower compared to, for example, the UK with a cumulative confirmed number of cases of 2.6%, Italy of 2.9%, and the USA of 4.7% (by December 2020) [11]. Incidence was also low with 2.5 new cases per 100 person-years. Results overall are comparable to what has been found for cystic fibrosis (CF). A French study of 7500 CF patients from 47 clinics found that 31 had tested positive for SARS-CoV-2 by June 2020, 0.4% of the study population [12]. Among the 31, 61% had been hospitalized and 13% (4/31) were in the ICU. [12]. For PCD, we found that most people who tested positive for SARS-COV-2 had mild symptoms, only 1 (8%) was hospitalised and none was in the ICU. The difference between the two studies might partly be explained by a higher degree of detection bias in the hospital-based French study, where mild infections not resulting in hospitalisations might have been missed by the physicians. This bias is less relevant, albeit not absent, for our participatory study with patients themselves reporting weekly on their health. In our study, only 45% had been tested for SARS-CoV-2 and it is possible that some participants had an undetected infection which would lead to an underestimation of the incidence. However, if a SARS-CoV-2 infection was missed, the participant most likely had mild symptoms. The reassuring results are probably partly explained by the careful shielding behaviour of our study participants – on average, 10% had not left their house in the past week and less than half had gone to school or work. But even so, the study suggests that with careful personal protection, people with PCD do not seem to have an increased risk of infection with SARS-COV-2, nor an especially severe disease course.

## Data Availability

Researchers who wish to use data should submit a concept sheet
describing the planned analysis and send it to Prof. Claudia Kuehni
. The concept sheet will be discussed with the
patient advisory group and if agreed, a partial dataset will be prepared by the
research team.

## Acknowledgements

We thank all participants and their families, and we thank the PCD support groups and physicians who have advertised the study. We thank our collaborators who helped set up the COVID-PCD study: Eugenie Collaud, University of Bern; Rebeca Mozun, University of Bern; Cristina Ardura, University of Bern; Yin Ting Lam, University of Bern; Christina Mallet, University of Bern; Helena Koppe, University of Bern; Dominique Rubi, University of Bern.

